# Comparison Canal Filling Ratio And Femoral Bone Density Change Between Wedge Taper And Anatomical Stem Design

**DOI:** 10.1101/2022.11.08.22282094

**Authors:** Patcharavit ploynumpon, Thakrit Chompoosang

## Abstract

**Purpose:** comparative the outcome of proximal femoral bone density change in follow-up x-ray film and proximal filling ratio of stem between anatomical and double taper wedge cementless stem design

**Methods:** post-operative follow film of up to 1 year of patients who had undergone Total hip arthroplasty between 2552-2563, which is match inclusion criteria, was obtained from the radiology department. The measurement of Canal filling ratio (Lesser trochanter, 2 cm above LT and 7 cm below LT) and Femoral bone density change using optimal densitometry method to compare between Anatomical and double wedge taper stem type.

**Result:** 92 patients,76% female, and 24% male, were match the inclusion criteria for this study. The mean age was 53.86±13.00 years old. The canal filling ratio in the double wedge taper group (Accolade II) was significantly higher than the anatomical stem group (ABGII) (p<0.001, p<0.001, p=0.013) in all levels of measurement. There were no significant differences between both types of the stem in femoral bone density change in zone 1,4. However. There were significant differences in femoral bone change, in which bone loss was higher in the anatomical stem group, in zone 7 (−25 VS −17, P= 0.010)

**Conclusion:** Double taper wedge stem design had a significantly higher canal filling ratio than the Anatomical stem at all levels and less femoral bone density loss in follow-up post-operative film at Zone 7. However, in zone 1,4, There was no significant difference in femoral bone density loss.

## Introduction

Total hip arthroplasty is one of the most common procedures preforms by orthopedic surgeons. As the number of surgery was raised, The stem design was one of the crucial factors distributed to the longevity of the overall prosthesis and the satisfaction of the patients. The cementless stem, invented in 1950, is one of the stem designs that provided Good results and long-term outcomes [1]. However, the early design was reported in several studies for early loosening and instability due to Proximal femoral osteopenia from the stress shielding effect [2]. Many modern stem designs were developed by promoting proximal engagement, by using HA porous coating and more fitting to the patient proximal femoral dense bone using a taper and anatomical designs, and decreasing the distal engagement of stem, using shorter stem design, which can lead to a decrease in proximal bone loss up to 14 % [3]. And there are many studies was shown that the revolution in stem design lead to less stem subsidence, less thigh pain, and less loosening [4-5]

However, there is no previous study comparing the progression of bone integration and proximal bone loss between Double wedge taper stem (Accolade 2 stem, Stryker) and anatomical stem (ABGII, Stryker). So our study aims to compare the difference in proximal femoral filling between both stem designs using immediate post-operative film and comparing proximal femoral bone loss using follow-up film x-ray. which is the result that will lead to a better choice of stem and a decrease in early complications and satisfaction with the total hip replacement operation.

## Material and method

### Study design

This study is a retrospective descriptive-cohort study of immediate and Post-operative follow film up to 1 year of Total hip arthroplasty surgery which is performed in Rajavithi hospital during 2017-2019 The Ethics Committee of Rajavithi hospital approved the research protocol in this study

### Patients

#### Inclusion Criteria

With permission from the radiology department of Rajavithi, Patients aged between 18-80 years old who received primary total hip arthroplasty using both types of stem who is performed between 2017-2019 and have follow-up films up to 1 year were included in this study

#### Exclusion Criteria

Patients under 18 years old, revision hip arthroplasty, patients with prior hip dysplasia, any post-operative complication, and patients who have follow-up film less than 1 year were excluded from the study.

#### Data Collection and measurement

After immediate and Post-operative follow film up to 1 year of the patients who had undergone Total hip arthroplasty were obtained from the radiology department, and the data was reviewed and analyzed. Immediate post-operative films were measured using canal filling ratio (Fig 1)[6], By measuring proximal femoral diameter and diameter of the stem in Anteroposterior view in 3 levels, lesser trochanter,2cm proximal to lesser trochanter and 6 cm distal to lesser trochanter by the orthopedic surgeon of adult hip and knee reconstruction.

**Fig 1:**
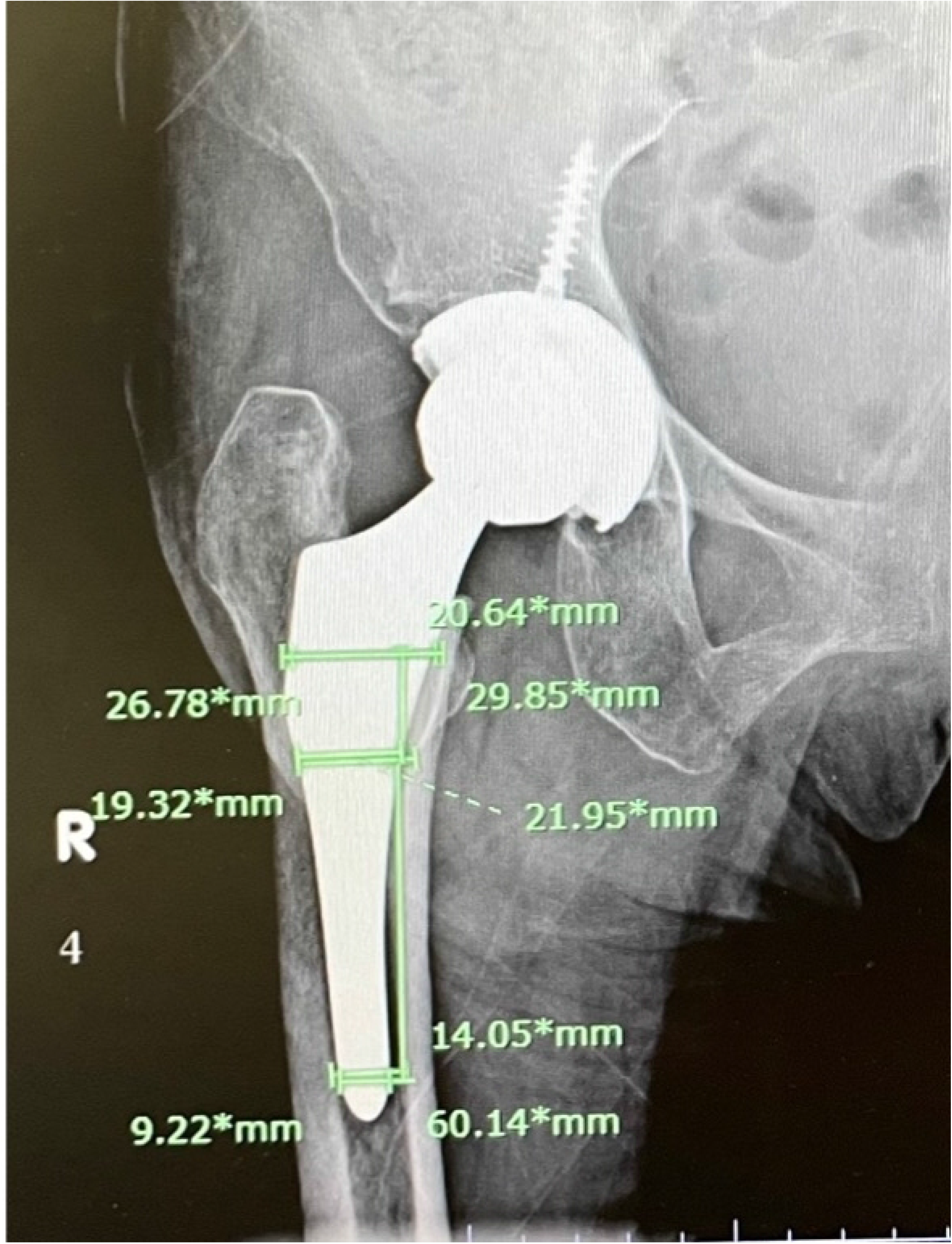
**Canal filling ratio measurement** measuring at lesser trochanter level2cm proximal to lesser trochanter and 6 cm distal to lesser trochanter

The follow-up film was analyzed for proximal femoral bone density change using optimal densitometry method [7] using digital optical image analysis program which is public domain, ImageJ for window, which measured bone change at zone 1,4,7 according to Gruen zone of fixation [8] (figure2) then all data was collected and recorded using Microsoft Excel

**Fig 2:**
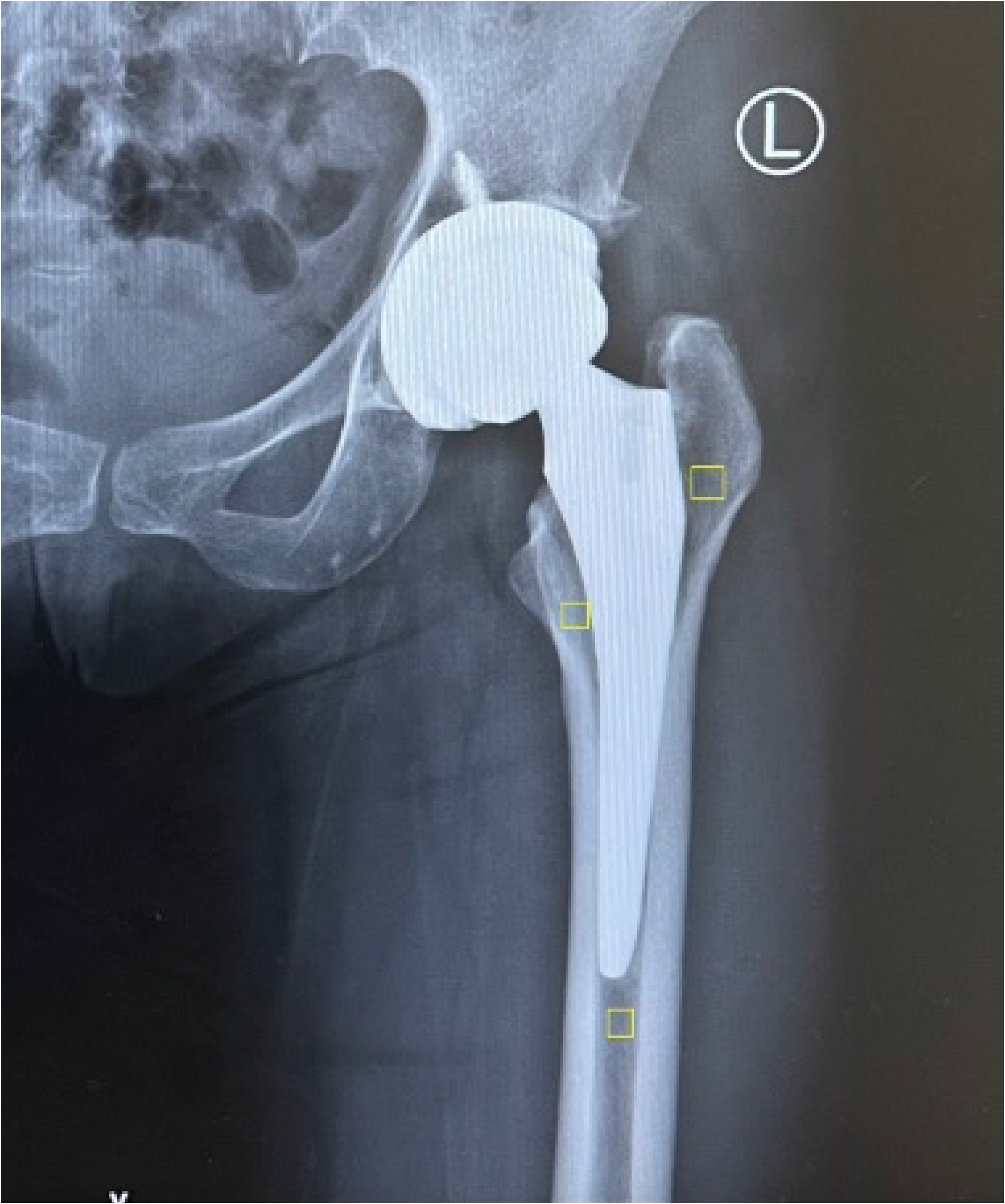
**Proximal femoral bone density measurement** using optimal densitometry method

### Statically analysis

Use descriptive statistics to describe the various patient characteristics of the sample. Use the number, percentage, Mean, Median, Standard deviation, minimum and maximum values. For Comparing category data using the Chi-square test. Comparing independent data such as both type of stem and femoral type by Paired T-test and using student T-test for dependent data such as comparing post-operative film. The level of significance was defined as a p-value <0.05. All statistical analyses were performed using SPSS Version 20

## Results

### Demographics data

92 patients were included in this study. 22 patients were male and 70 were female. Mean age 53.86±13.00 years old. 34 patients were in undergone anatomical stem (ABGII, Stryker) group and 58 were in undergone double wedge taper stem (Accolade II, stryker) group

When comparing the canal-filling ratio between both stems, the double wedge taper stem had a significantly higher canal-filling ratio than the anatomical stem group at all 3 measurement levels (p<0.001, p<0.001, p=0.013) as the result was shown in Table 2.

**Table 2:**
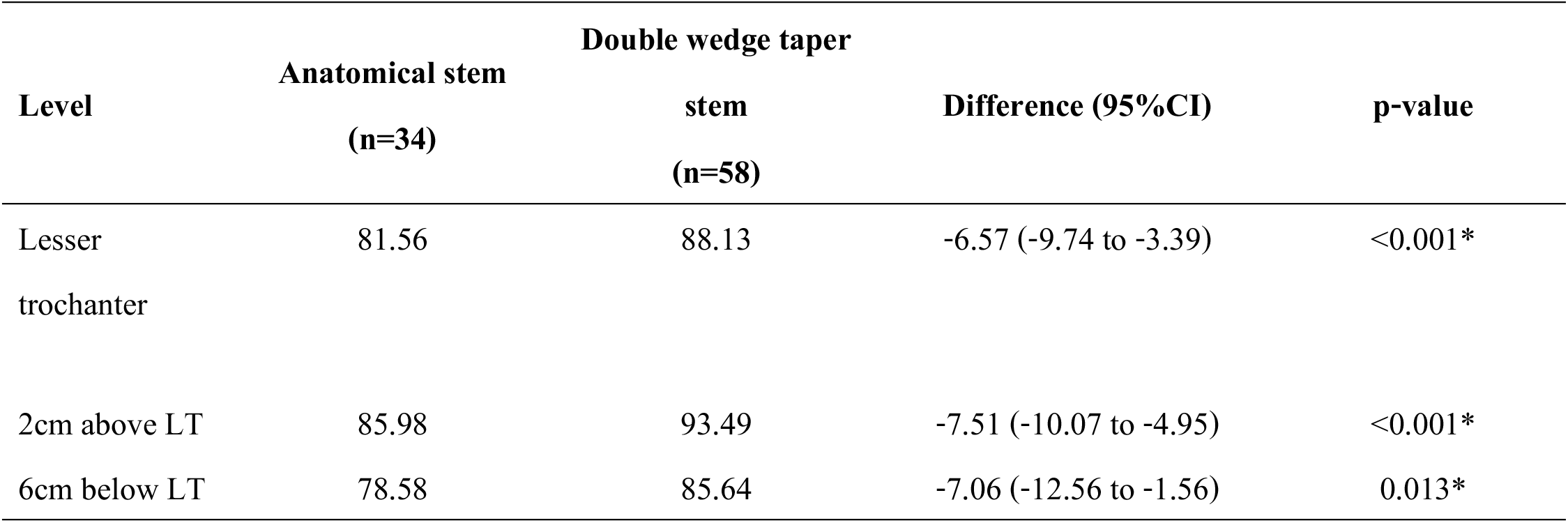
Comparison of canal filling ratio between 2 types of stem.

| Level | Anatomical stem | Double wedge taper | Difference (95%CI) | p-value |
| --- | --- | --- | --- | --- |
|  | (n=34) | stem<br>(n=58) |  |  |
| Lesser trochanter | 81.56 | 88.13 | -6.57 (-9.74 to -3.39) | <0.001* |
| 2cm above LT | 85.98 | 93.49 | -7.51 (-10.07 to -4.95) | <0.001* |
| 6cm below LT | 78.58 | 85.64 | -7.06 (-12.56 to -1.56) | 0.013* |

The post-operative film was analyzed in each of the stem designs as shown in Table 3. both of the stems showed a femoral proximal bone loss from the baseline and every time point.

**Table 3.**
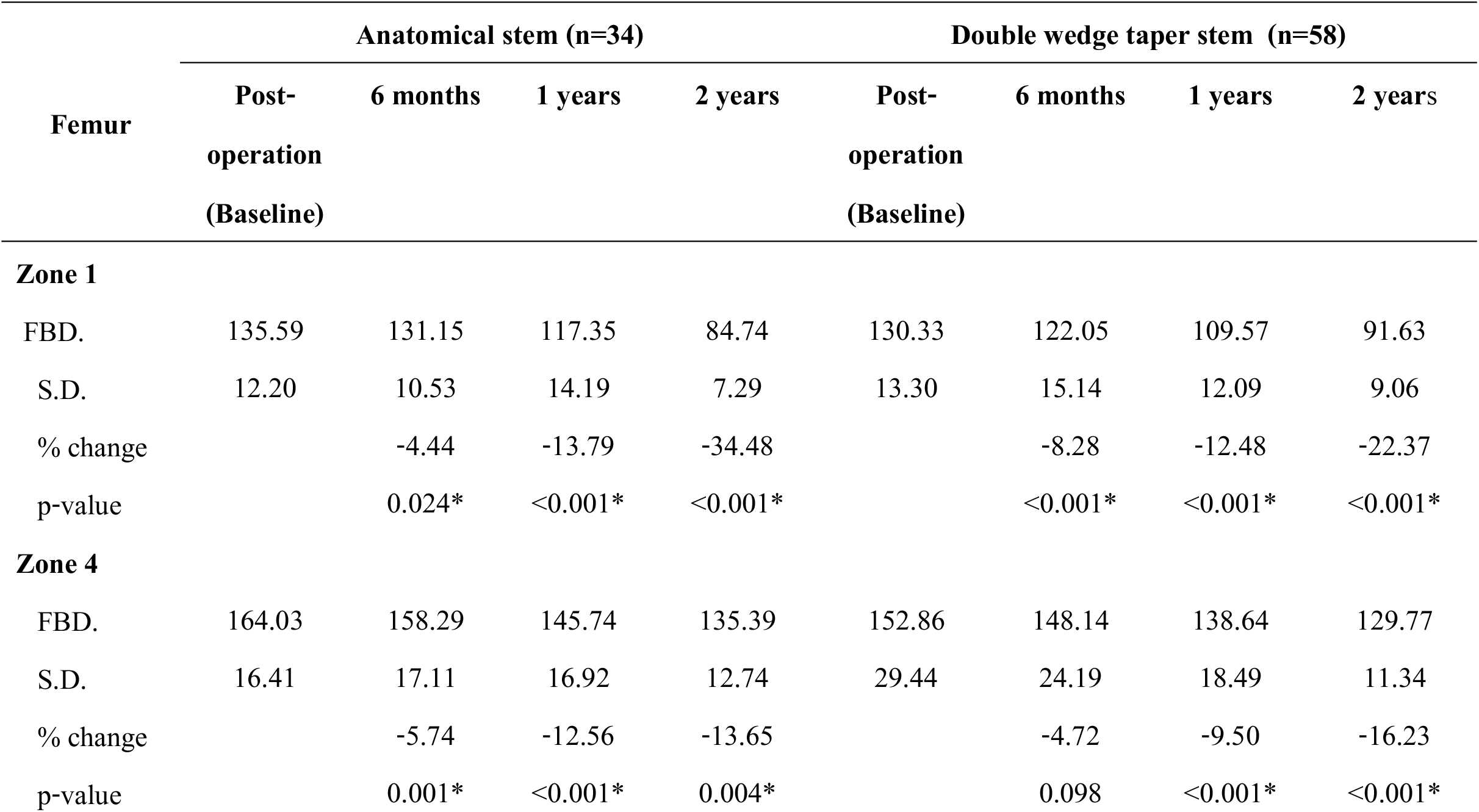
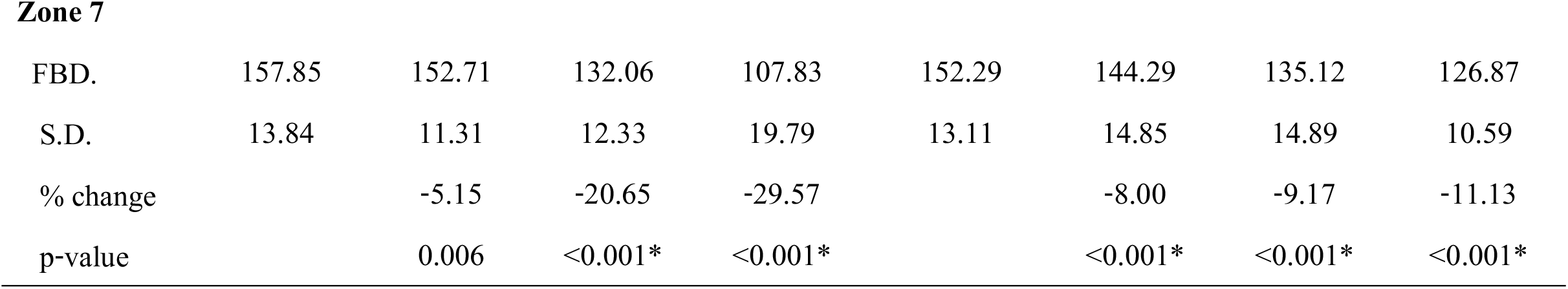
**Femoral bone density change in each stem type** in Gruen zone 1,4,7

#### 1. Proximal bone density change in anatomical stem

At 6 months postoperatively, There was a significant difference in femoral bone loss at zone 1,4,7 (p=0.024, p<0.001, p=0.006 respectively). The most femoral bone loss is at zone 4 (5.74)

At 1 year postoperatively, There was a significant difference in femoral bone loss at zone 1,4,7 (p<0.001). The most femoral bone loss is at zone 7 (20.65)

At 2 years postoperatively, There was a significant difference in femoral bone loss at zone 1,4,7 ((p<0.001, p=0.004, p<0.001 respectively). The most femoral bone loss is at zone 1 (34.48)

#### 2. Proximal bone density change in double wedge taper stem

At 6 months postoperatively, There was a significant difference in femoral bone loss at zone 1,4,7 (p<0.001). The most femoral bone loss is at zone 1 (8.28)

At 1 year postoperatively, There was a significant difference in femoral bone loss at zone 1,4,7 (p<0.001). The most femoral bone loss is at zone 1 (12.48)

At 2 years postoperatively, There was a significant difference in femoral bone loss at zone 1,4,7 (p<0.001,). The most femoral bone loss is at zone 1 (22.37)

Comparing proximal femoral bone loss between both stem designs, the double wedge-taper stem showed significantly less proximal femoral bone loss in Gruen zone 7(Fig 5). However, There were no significant differences in proximal femoral bone loss in Gruen zone 1,4 as shown in (Fig3-4) and Table 4.

**Fig 3.**
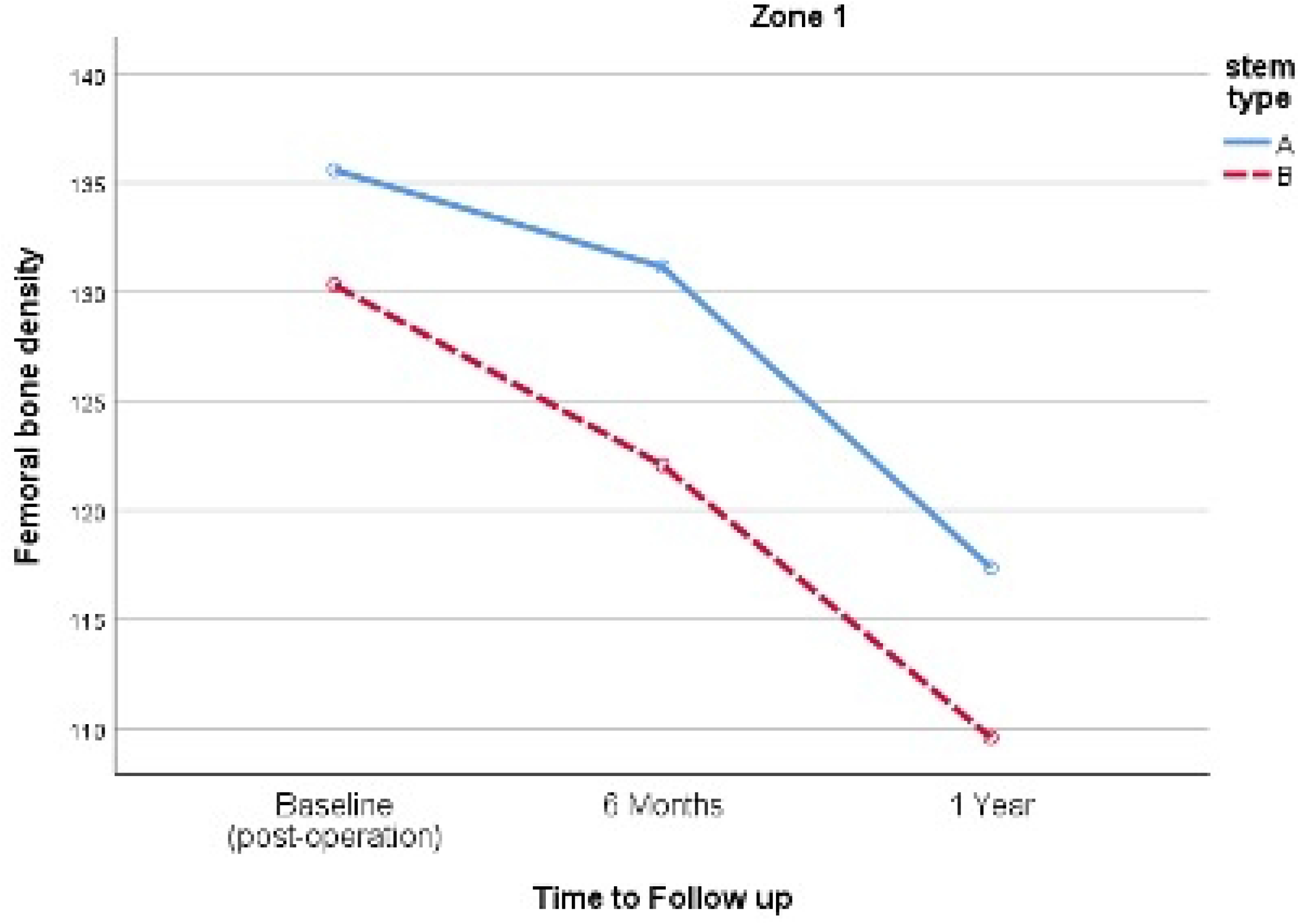
Comparing proximal femoral bone density change in Gruen Zone 1 of both stem (A=anatomical stem, B=double wedge taper stem)

**Fig 4.**
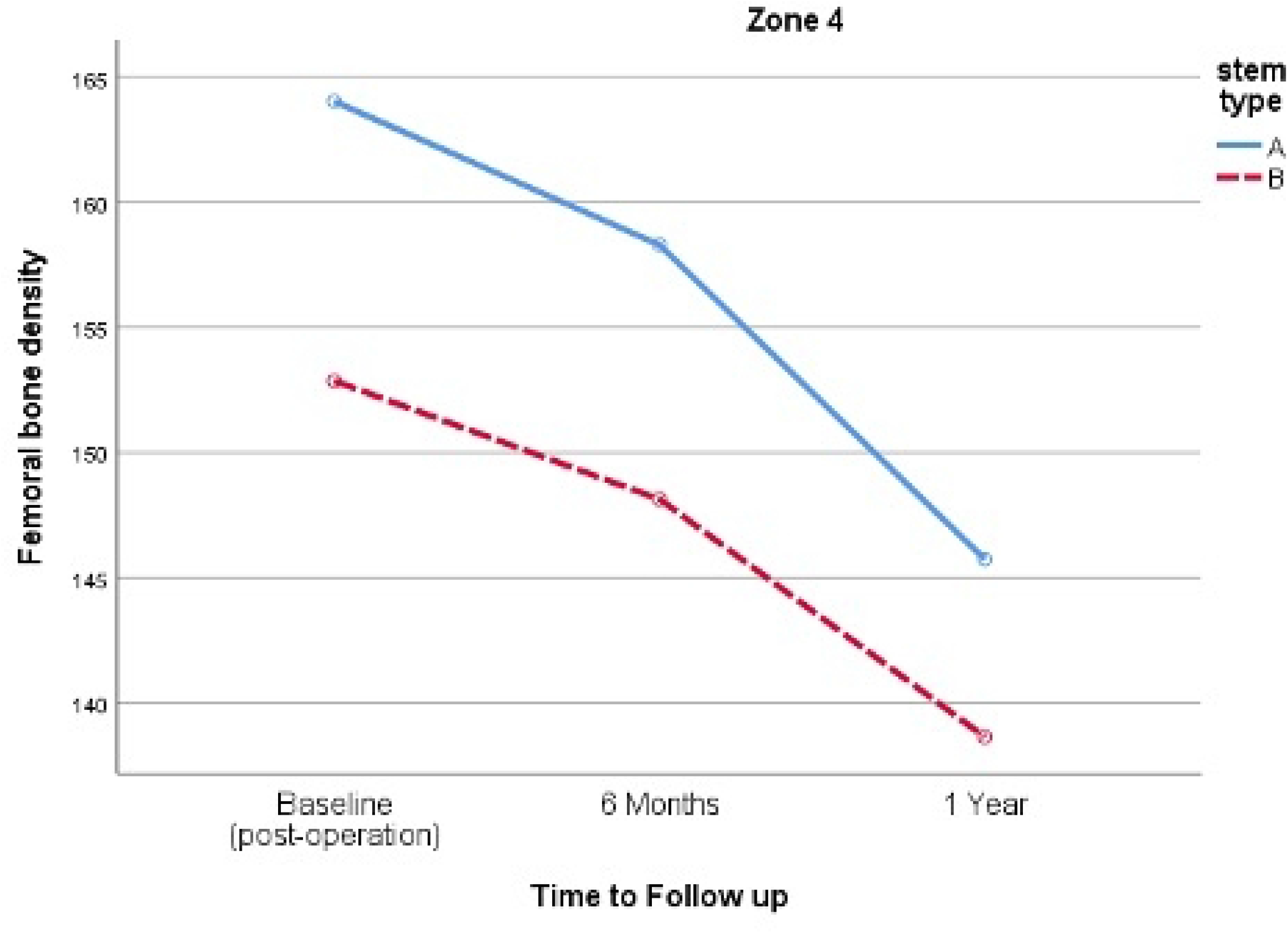
Comparing proximal femoral bone density change in Gruen Zone 4 of both stem (A=anatomical stem, B=double wedge taper stem)

**Table 4.** **Comparison of Femoral bone density change in each zone in both types of stem** The table shows a significant difference in femoral bone density change only in zone 7 between both stems (P=0.01)

| Femoral bone density | Stem Type |  | p-value |
| --- | --- | --- | --- |
|  | Anatomical stem | Double wedge |  |
|  | (n=34) | taper stem (n=58) |  |
| Zone 1 |  |  |  |
| Post-operation (Baseline) | 135.59±12.20 | 130.33±13.30 | 0.062 |
| 6 months | 131.15±10.53 | 122.05±15.14 | 0.003* |
| 1 years | 117.35±14.19 | 109.57±12.09 | 0.006* |
| 2 years | 84.74±7.29 | 91.63±9.06 | 0.004* |
| Change (1 years – post-operative) | -18.24±20.48 | -20.76±9.36 | 0.501 |
| Zone 4 |  |  |  |
| Post-operation (Baseline) | 164.03±16.41 | 152.86±29.44 | 0.022* |
| 6 months | 158.29±17.11 | 148.14±24.19 | 0.021* |
| 1 years | 145.74±16.92 | 138.64±18.49 | 0.070 |
| 2 years | 135.39±12.74 | 129.77±11.34 | 0.096 |
| Change (1 years – post-operative) | -18.29±14.79 | -14.22±24.22 | 0.320 |
| Zone 7 |  |  |  |
| Post-operation (Baseline) | 157.85±13.84 | 152.29±13.11 | 0.058 |
| 6 months | 152.71±11.31 | 144.29±14.85 | 0.005* |
| 1 years | 132.06±12.33 | 135.12±14.89 | 0.314 |
| 2 years | 107.83±19.79 | 126.87±10.59 | <0.001* |
| Change (1 years – post-operative) | -25.79±15.85 | -17.17±13.23 | 0.010* |

**Fig. 5.**
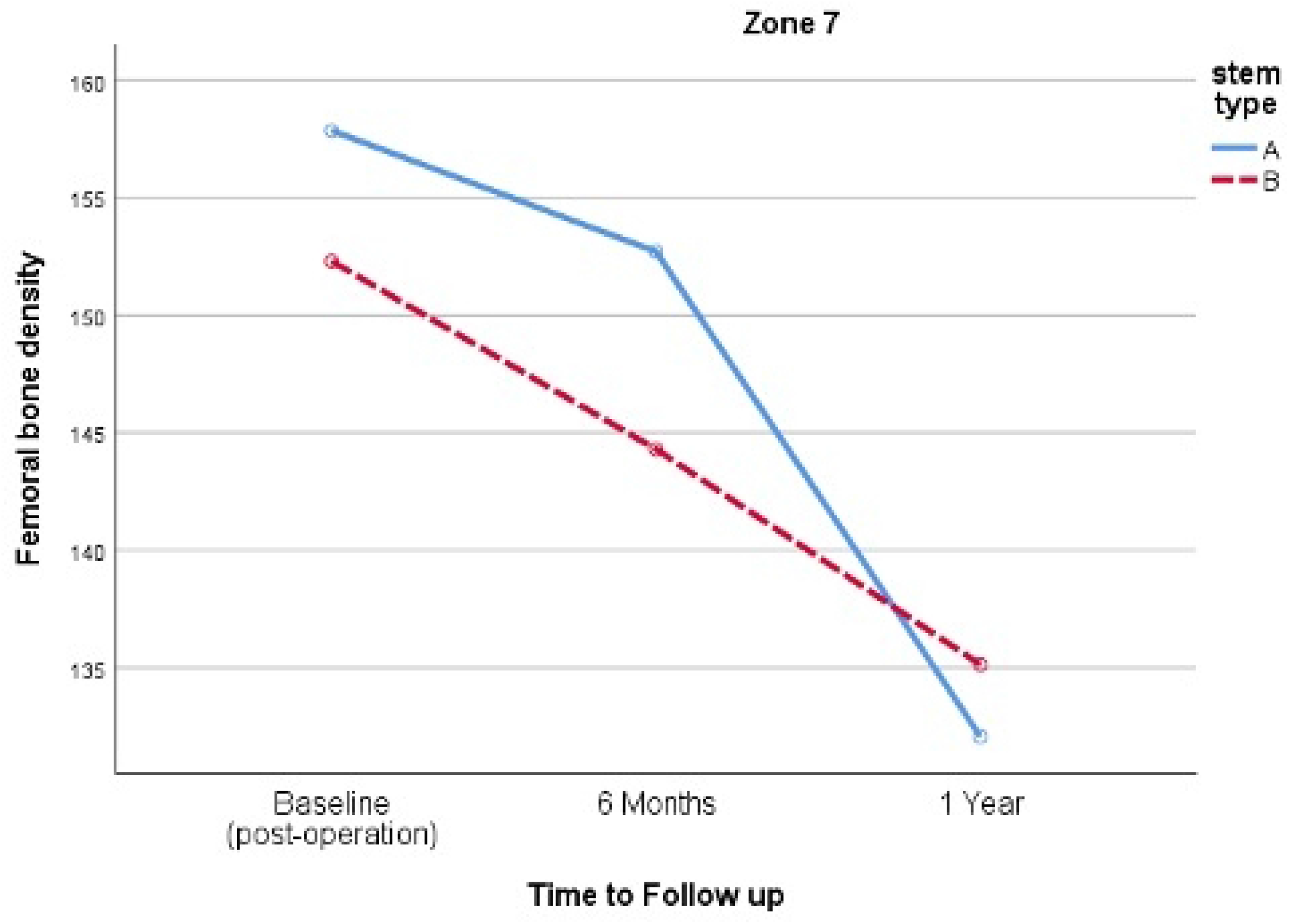
Comparing proximal femoral bone density change in Gruen Zone 7 of both stem (A=anatomical stem, B=double wedge taper stem)

## Discussion

Total hip arthroplasty is one of the most common procedure was performed by orthopedic surgeons. Cementless total hip arthroplasty is one of the popular procedures to be performed, especially for younger patients [9], which can achieve a good long-term outcome, Although There were reported proximal femoral osteopenia and early aseptic loosening in early design [4] due to stress shielding effect and proximal micromotion of the stem. Later, The cementless stem was developed by decreasing distal filling, increasing proximal filling, and improving proximal coating to achieve better osteointegration

In our study, we found a significantly higher canal filling ratio in the double wedge taper stem than in the anatomical stem group at all levels (LT,2 cm above LT, 6 cm below LT) and There was significantly higher femoral bone density loss at zone 7 in the anatomical stem than double taper wedge stem (−25 VS −17, P= 0.010), however, there was no significant femoral bone density loss in zone 1,4 .which is corresponding with a previous study of Cooper et al. found that increase distal filling ratio which leads to less osteointegration [10]

The strength of this study was that it was conducted by a single surgeon and a single center which minimized the confounding factor from surgical technique and post-operative care of the patients.

The limitation of our study is that it was retrospective in design, making it difficult to acquire data on some patients. Another limitation was the short follow-up time frame, which could lead to underestimating the complication rate and loss of femoral bone density.

## Conclusion

Double taper wedge stem design had a significantly higher canal filling ratio than the Anatomical stem at all levels and less femoral bone density loss in follow-up post-operative film at Zone 7. However, in zone 1,4, There were no significant differences in femoral bone density loss.

## Data Availability

All relevant data are within the manuscript and its Supporting Information files

